# Adjusting for reporting lags in remote regions to improve malaria surveillance: Guyana as a case study

**DOI:** 10.1101/2020.09.08.20178756

**Authors:** Tigist F. Menkir, Horace Cox, Canelle Poirier, Melanie Saul, Sharon Jones-Weekes, Collette Clementson, Pablo M. de Salazar, Mauricio Santillana, Caroline O. Buckee

## Abstract

Time lags in reporting to national surveillance systems represent a major barrier for the control of infectious diseases, preventing timely decision making and resource allocation. This issue is particularly acute for infectious diseases like malaria, which often impact rural and remote communities the hardest. In Guyana, a country located in South America, poor connectivity among remote malaria-endemic regions hampers surveillance efforts, making reporting delays a key challenge for elimination. Here, we analyze 13 years of malaria surveillance data, identifying key correlates of time lags between clinical cases occurring and being added to the central data system. We develop nowcasting methods that use historical patterns of reporting delays to estimate occurred-but-not-reported monthly malaria cases. To assess their performance, we implemented them retrospectively, using only information that would have been available at the time of estimation, and found that they substantially enhanced the estimates of malaria cases. Specifically, we found that the best performing models achieved up to two- to four-fold improvements in accuracy (or error reduction) over known cases in selected regions. Our approach provides a simple, generalizable tool to improve malaria surveillance in endemic countries.

## Introduction

Malaria continues to be one of the most important causes of morbidity and mortality globally - with 405,000 deaths estimated to have occured in 2018 - particularly in poor and rural populations.^1^ National malaria surveillance programs depend on timely reporting rates to rapidly identify signals of increasing malaria activity and potential outbreaks to ensure that control resources are optimally and most efficiently deployed.^2^ However, major barriers to timely surveillance in endemic countries - particularly those in which malaria transmission is concentrated in the most remote, hardest to reach populations - include limited connectivity, modes of digitisation and other context-specific factors that undermine the ability to update surveillance records without significant delays.^3^ While infrastructure development is critical to improving the timeliness of case reporting, methodological approaches to adjust for known reporting lags are urgently needed until data systems improve.

One such endemic context is Guyana, located in the northern coast of South America and bordering Venezuela, Suriname, and Brazil; one of 21 malaria endemic countries in the Americas.^4^ Guyana has a relatively small, sparse population of ~800,000 people^5^, with highly seasonal malaria endemicity concentrated in rainforest regions with low accessibility, often only reachable by small plane or boat. While confirmed cases have steeply declined from 2015 to 2017 in South America overall, Guyana has experienced a near 40% escalation in confirmed cases during that period, with 13,936 cases confirmed in 2017.^4^ More than half of infections are caused by *Plasmodium vivax;* however, the second most common (44%) infecting species, *P. falciparum*, is tied to more severe outcomes and has been implicated in recent reports of resistance to Artemisinin drugs.^6–8^ The recent growth in the number of confirmed case counts overall is partially driven by increased importation from surrounding areas, such as Venezuela, linked with migration from social and political crises and to promising employment opportunities in mining.^4,9^ In response to the elevated burden of malaria in the nation, the government has boosted investment in malaria prevention, under the direction of the Vector Control Services (VCS).^4,10^

Like many national malaria control programs, one of the priority areas of VCS is surveillance, where regional health offices and the central office in the capital of Georgetown engage in a complex surveillance cascade. Health facilities in endemic regions are required to send their daily case registers for all malaria patients to their corresponding regional health office for processing, before being submitted to the central office in the capital of Georgetown, while health facilities in non-endemic regions can simply send reports to Georgetown.^11^ However, we find that many health facilities in endemic areas opt to send their reports directly to Georgetown or through alternative routes, due to issues in road access and other challenges, often resulting in delayed or missed reports (*personal communication*). Furthermore, while the data is intended to be documented in the system in near real-time, with summary reports sent from regions to the central office via USBs, physical copies of individual case registers from facilities are sometimes delivered to the central office, leading to additional bottlenecks in reporting (*personal communication*). Together, these challenges contribute to the number of cases recorded in the central database at the end of the month potentially failing to reflect the true burden of disease at a given time, with cases typically reported more than a month after they occur (See Figure 1). These issues are not only common for malaria control and surveillance programs, but for many other epidemiological reporting systems, hindering the ability of public health programs to implement appropriate and optimal measures for control.^3,12–16^ Modeling efforts to translate available reporting data to more meaningful and accurate measures of real-time incidence could help address some of these challenges.

**Figure 1:**
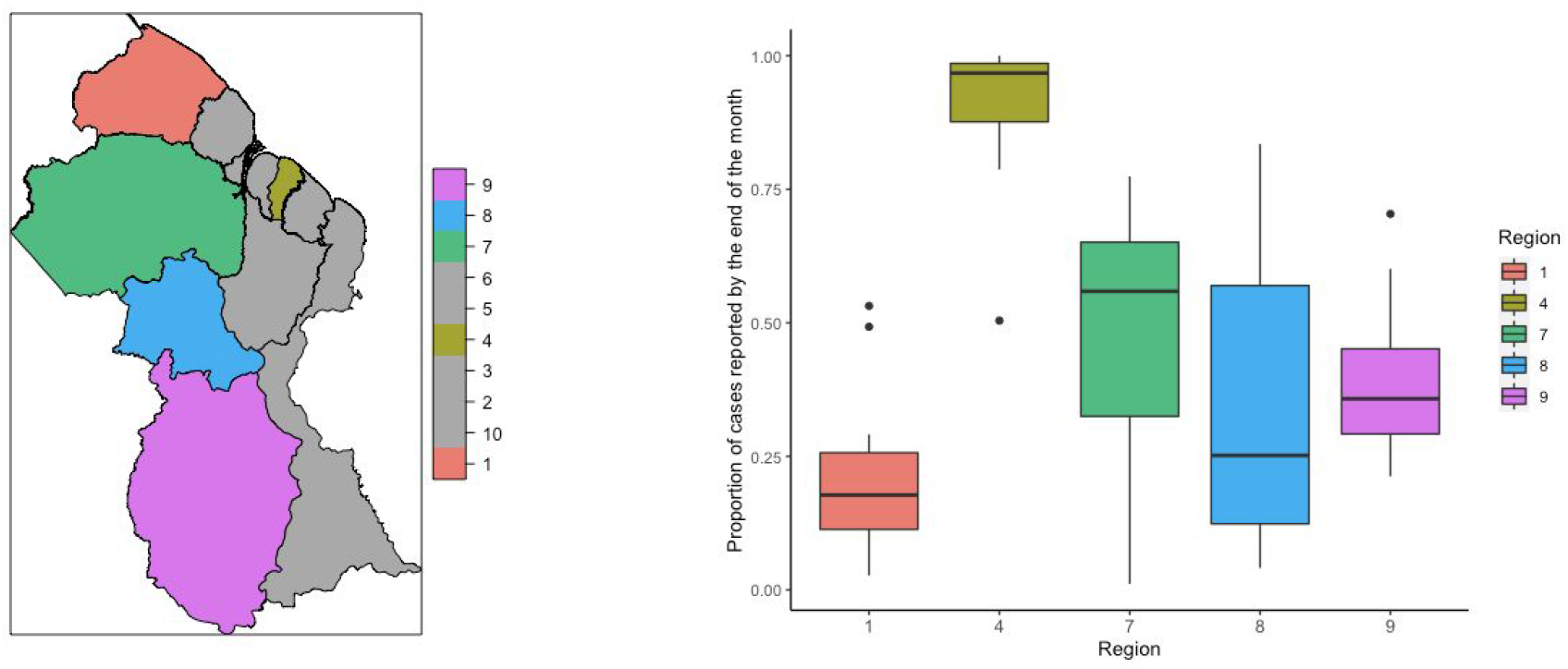
Map of malaria endemic regions and region 4 (left) and boxplot reporting the proportion of annually aggregated cases from 2006-2019 reported by the end of the month for each region, with colors corresponding to the regions depicted in the map (right)

A number of analytical approaches aimed at improving estimates through “nowcasting” have been developed to account for reporting delays (frequently referred to as “backfill”). The most widely used are Bayesian estimation and regularized regressions, which take advantage of the fact that the timing of delays may be relatively predictable. The Bayesian method encapsulates both delay trends and epidemiological trends to estimate disease occurrence.^14,15,17^ Extensions of this approach, proposed by Bastos et al. and Rotejanaprasert et al., additionally accommodate spatial trends in transmission and delay dynamics.^17,14^ Relatedly, Reich et al. use a generalized additive model to estimate Dengue Hemorrhagic Fever cases in Thailand, modeling incidence at each time point in a given province as a function of its preceding reported cases and of previous reported cases from provinces with similar activity.^16^ One limitation of the Bayesian methods is that they do not focus on providing more interpretable measures to guide actionable surveillance efforts, such as direct point estimates for predicted case counts.^18^ Penalized regression methods, such as the ARGO, Net, and ARGONet models proposed by Santillana, Lu, and others have also been used to incorporate a diversity of case reporting and alternative data sources, to learn from previous time trends in delays and transmission, *and* the spatial dependencies in these trends, to predict rates of influenza-like illness.^19^ However, this class of models is only well-suited for contexts in which these varied digital sources are readily available and easily integrated into routine model training, testing, and validation efforts.^19^

To address challenges in reporting delays for malaria surveillance in Guyana, we developed nowcasting approaches to estimate monthly cases of malaria in each endemic region, using data from the central VCS database from 2006 to 2019 to develop and test our models. We show that proximity to high-risk vulnerable populations, namely mining sites and Amerindian populations, is associated with reporting delays and that delays are relatively consistent over time. Importantly, our predictions provide extensive improvements in surveillance capacity for remote areas. Thus, we illustrate that nowcasting offers a general, tractable approach for improving decision-making for malaria control programs in countries that have significant reporting delays.

## Methods

We had access to daily case register data from 2006 to 2019 documented at health facilities in each of Guyana’s ten regions and compiled at the central office in Georgetown (region 4). Delays are defined as the days elapsing from when a patient is registered as a case at a health facility and when the patient is documented at the central VCS database.

We assessed the potential influence of rainfall on reporting delays by analyzing precipitation data sourced from East Anglia University’s Climate Research Unit’s gridded dataset repository. Monthly total precipitation data was extracted for each region by year.^20^ Locations of second-level administrative regions, or neighborhood democratic councils (NDCs) (n=116) were identified through a shapefile of NDCs from GADM version 1 (accessed through DIVA-GIS).^21^ We defined NDCs, rather than regions, as our spatial unit of analysis to ensure adequate statistical power, and aggregated median delays across years by locality for each NDC. Connectivity was captured through measures of accessibility (defined as travel times to the “nearest urban centre”) for each region in 2015 estimated in Weiss et al.^22^ A shapefile of mapped mines was drawn from the 2005 U.S. Geological Survey Mineral resources data system, respectively.^23^ Finally, a shapefile of Amerindian settlements was obtained from the LandMark Global Platform of Indigenous and Community Lands through the GuyNode Spatial Data portal.^24^ For all analyses, we excluded the 5.7% of observations with implausible recorded date ranges, i.e. individuals whose smear was examined in a health facility following their documentation in the central office in Georgetown.

### Descriptive analysis

We first assessed the stationarity of each region’s monthly delay distributions from 2006 to 2019 through an Augmented Dickey-Fuller test. We further examined potential synchronicities between regions’ monthly delay distributions, using the mean cross-correlation coefficient between the regional time series for monthly delays from 2006 to 2019.

For each region, we additionally estimated (a) the Pearson correlation between median reporting delays and pairwise region connectivity in 2015, and (b) the Pearson correlation and cross-correlations between monthly total precipitation levels and median delays from 2006-2019. We focused our analysis on case register data from region 4, where many malaria patients come for treatment but there is no local transmission, and from malaria-endemic regions 1, 7, 8, and 9 (see Figure 1 for map).

We used a global Moran’s I index^25^, to evaluate if delays across NDCs from 2017-2019 (the most recent three years of data) across all ten regions were spatially autocorrelated. This measure estimates whether, and to what extent, observations in the same neighborhood --defined using a range of possible criteria-- share “features” which would indicate spatial autocorrelation.^25,26^ We further used the Getis-ord statistic, which compares the sum of characteristics in each neighborhood to their overall mean^26,27^, to identify local clusters of NDCs with higher reporting delays. To pinpoint the potential correlates of the observed spatial patterns in delays, we computed the local bivariate Moran’s I statistic for NDC-level delays and density of Amerindian settlements and for NDC-level delays and density of mines. We chose not to compute the local bivariate Moran’s I for NDC-level delays and connectivity, which we expect to exhibit a particularly strong spatial relationship with delays, given that we were restricted to data on connectivity at the scale of regions. For all spatial analyses, we omitted the 2.9% of patients whose localities could not be geocoded. Spatial weights were defined using the queen’s contiguity criterion of order=1. All significance maps for local autocorrelation tests that we provide report FDR-adjusted p-values.

### Nowcasting methods

We used the following two approaches to estimate revised (i.e. eventually reported) monthly malaria case counts in regions 1,4,7,8, and 9. In all of the proposed approaches, our real-time malaria case count estimations were produced in a strictly out-of-sample fashion, that is, only information that would have been available at the time of estimation was used to train our models. The first out-of-sample case count estimate for all regions was produced for January of 2007 using historical information available at the time (training set time period) that consisted of data from the previous 12 months (within 2006). Subsequent estimates were produced by dynamically training the models below, such that as more information became available the training set consisted of a new set of (12-month long) observations. This approach was chosen as a better way to capture the potential time-evolving nature of reporting delays.

#### Synchronous data imputation model (DIM)

For each region i, we use an elastic net penalized regression, i.e. an ordinary least squares regression (OLS) subject to a combined L1- and L2- penalty^28^, to estimate the revised case count for a given month t. The model was fit to data on the number of cases occurring in months t to t-12 that were known by the end of month t and takes the following form (1), where 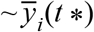 denotes the number of cases occurring in each month t* known by the end of month t:

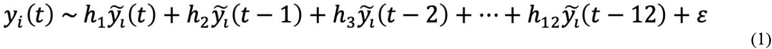

### Data inputs and method of training and testing the simple data imputation model

A matrix of case counts from each of the twelve months prior to time and including time t (the most up to date case count) that were known by the end of time t was used to dynamically train and test the data imputation model in order to predict the number of revised cases for t. Our models were assessed in their ability to accurately predict the revised case counts, only available at time t+m, where m was typically more than a month later, and were compared to the case counts available at time t. We computed a 3-fold cross validation within each twelve month window in order to identify the tuning parameters that resulted in the lowest mean square error and used these parameters for the subsequent (unseen) monthly prediction. In other words, training of our models was conducted using strictly data that would have been available at the time of prediction. Given the size of the training set was 12 observations, we chose a small number of folds, n = 3 for our cross-validation implementation. We experimented with different number of folds (3, 5, or 10) for cross-validation to explore whether this choice would have an impact on the quality of our predictions and found that these additional choices led to qualitatively similar results, so we report the results of the most cost-efficient approach in the manuscript.

#### Synchronous network models (NM)

For each region i, an elastic net regression estimating the revised case count for a given month t was fit to data on the number of cases occurring in months t to t-12 that were known by the end of month t, the number of cases occurring in months t-1 to t-12 that were known by the end of the month t in all other regions j ≠ i, and the total precipitation level in region i at month t, a hypothesized cofactor of reporting activity. We also ran an elastic net regression which additionally takes as inputs the predicted number of cases in month t for all other regions j from the previous data imputation models.

The first network model takes the following form (2), where θ(*t*) denotes the total precipitation level for region i at time t.

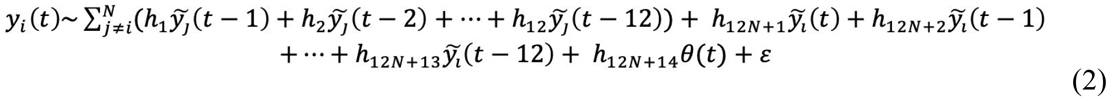

The second network model takes the following form (3), where *y_j_*(*t*) denotes the predicted case counts for region j at time t, estimated from the data imputation model for region j (1)

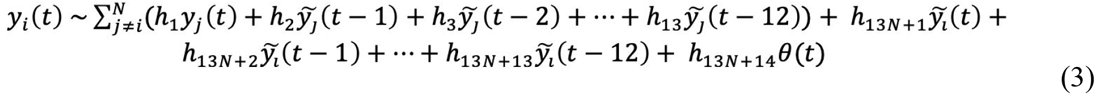

### Data inputs and method of training and testing the two network models

A matrix of case counts from each of the twelve months prior to and including t that were known by the end of t for region i, case counts from each of the twelve months prior to t that were known by the end of t for all other regions j, and total precipitation levels for region i in month t, was used to dynamically train and test NM1 in order to predict the number of revised cases for t for region i.

A matrix of case counts from each of the twelve months prior to and including t that were known by the end of t for region i, case counts from each of the twelve months prior to t that were known by the end of t for all other regions j, DIM-predicted case counts in t for all other regions j, and total precipitation levels for region i in t was used to dynamically train and test NM2 in order to predict the number of revised cases for t for region i.

We replicate the same procedure as in the data imputation model to test and train the two network models.

All data pre-processing, a-spatial and nowcasting analyses were conducted in R version 1.2.1335.^29^ All spatial analyses were conducted in ArcMap version 10.6.1 and GeoDa.^30,31^

## Results

### 1. Descriptive Analyses

Between 2006 and 2019, malaria cases were entered in the central database a median of 32 days after they were registered at local clinics, with a marked right skew in the distribution of delays, for all regions combined (sd=52.7). Figure 1 illustrates how these delays occur in different regions (see Figure S1 for full empirical density functions), and highlights the considerable heterogeneity in the extent of reporting delays observed between them, with region 1 ranking lowest in the proportion of annually aggregated cases from 2006-2019 reported by the end of the month (median=0.18, 95% CI=0.11,0.26) and region 4, where the capital is located, ranking highest, reporting nearly all of its cases by the end of the month (median=0.97, 95% CI=0.88,0.99). Neighboring regions 7, 8 and 9 additionally capture a low fraction of cases by the end of the month (median=0.56, 95%CI = 0.32,0.65, median=0.25, 95% CI=0.12,0.57; median=0.36, 95% CI=0.29,0.45, respectively).

Monthly total precipitation was not correlated with monthly median delays between 2006 and 2019 for any of the regions (p-value=0.89, p-value=0.80, p-value=0.41, p-value=0.62, p-value=0.84, for regions 1,4,7,8 and 9, respectively). While we expect that the differences between region 4 and the other regions in reporting delays are likely due to the remoteness of regions 1, 7, 8, and 9, the estimated association between region-specific connectivity and median delays in 2015 was modest and non-significant (r=-0.13, p-value=0.71). Total precipitation and monthly delays for all endemic regions reported no significant cross-correlations within a meaningful range in lags.

We observed significant spatial autocorrelation in median reporting delays by NDC for the most recent years of data (Global Moran’s I=0.496,p=0.001). NDCs characterized by long delays were found in close proximity to NDCs with similarly high delays and vice versa (Figure 2 and Supplementary Figure S3). Significant clusters of NDCs marked by higher delays were most concentrated in regions 1 and 7, while clusters of NDCs marked by lower delays were most concentrated in region 4 and 6 (Figure 2 and Supplementary Figure S3). Delays were significantly positively spatially correlated with density of mines by NDC, with the bivariate analysis highlighting areas with NDCs characterized by elevated delays surrounded by a greater density of mines (largely concentrated in regions 1 and 7) (Figure 2 and Supplementary Figure S3) (bivariate Moran’s I=0.590, p=0.001). We also found evidence for a significant positive bivariate spatial association between reporting delays and density of Amerindian settlements, revealing areas with NDCs characterized by higher delays surrounded by a greater density of Amerindian settlements, mainly concentrated in regions 1, 8 and 9 (Figure 2 and Supplementary Figure S3) (bivariate Moran’s I=0.587, p=0.001). Both clustering maps signal NDCs within region 1 marked by higher reporting delays and a relatively increased presence of Amerindian settlements and mines. Note that NDCs showing the inverse of this relationship, i.e. lower delays surrounded by a higher density of Amerindian settlements or mines, were located in regions 6 and 10, where malaria is not endemic (Supplementary Figure S3).

**Figure 2:**
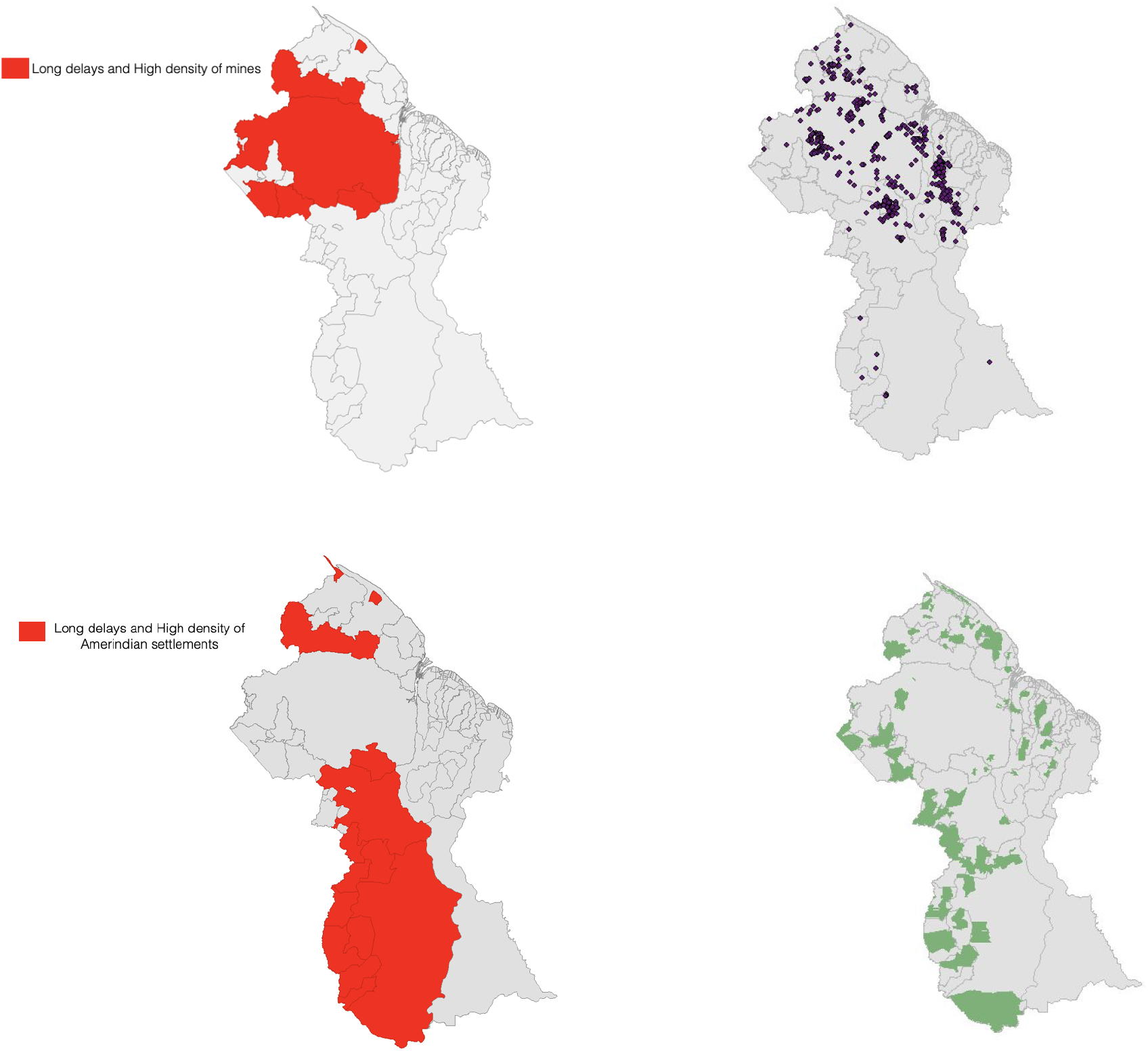
Local bivariate Moran’s I clustering maps of aggregated delays and density of mines, by NDC, and aggregated delays and density of Amerindian settlements, by NDC (top left and bottom left). For visual purposes, we show only high-high clusters of areas reporting at higher delays with a greater density of mines or Amerindian settlements. Full bivariate cluster and significance maps can be found in Supplementary Appendix, Figure S4. Map of mining sites and Amerindian areas (top right and bottom right)

We found weak evidence for overall synchronicity in delay distributions across regions, with an estimated mean cross-correlation between the regional monthly delay distributions equal to 0.24 (95% CI=0.20,0.27) and to 0.25 (95% CI=0.18,0.43) when excluding region 9, due to low case numbers. Overall, these minimally correlated delay distributions may reflect varying seasonality in transmission between regions. Finally, there was significant evidence for stationary of the time series of monthly delay distributions for all regions (from 2006-2019) (p-value=0.01 for all six regions). These findings support the use of data imputation models, which rely on previous trends in delays from previous years in a given region to inform ongoing predictions.

### 2. Nowcasting Results

#### A. Data Imputation Models (DIM)

The model based on region 4, where the capital Georgetown is located, yielded the best estimate of monthly cases from 2006 to 2019 (rRMSE=0.0791). DIM model predictions coincided with the two highest peaks in true cases and generally reflected trends through time, despite substantially underestimating a moderate peak in late 2012 and generally overshooting revised cases in late 2013 (Figure 3). Given that the number of cases known by the end of each month in region 4 closely approximates the number of revised cases, this approach may be of limited practical utility. Due to the high accuracy of region 4 DIM, we chose not to implement network models for this region.

**Figure 3:**
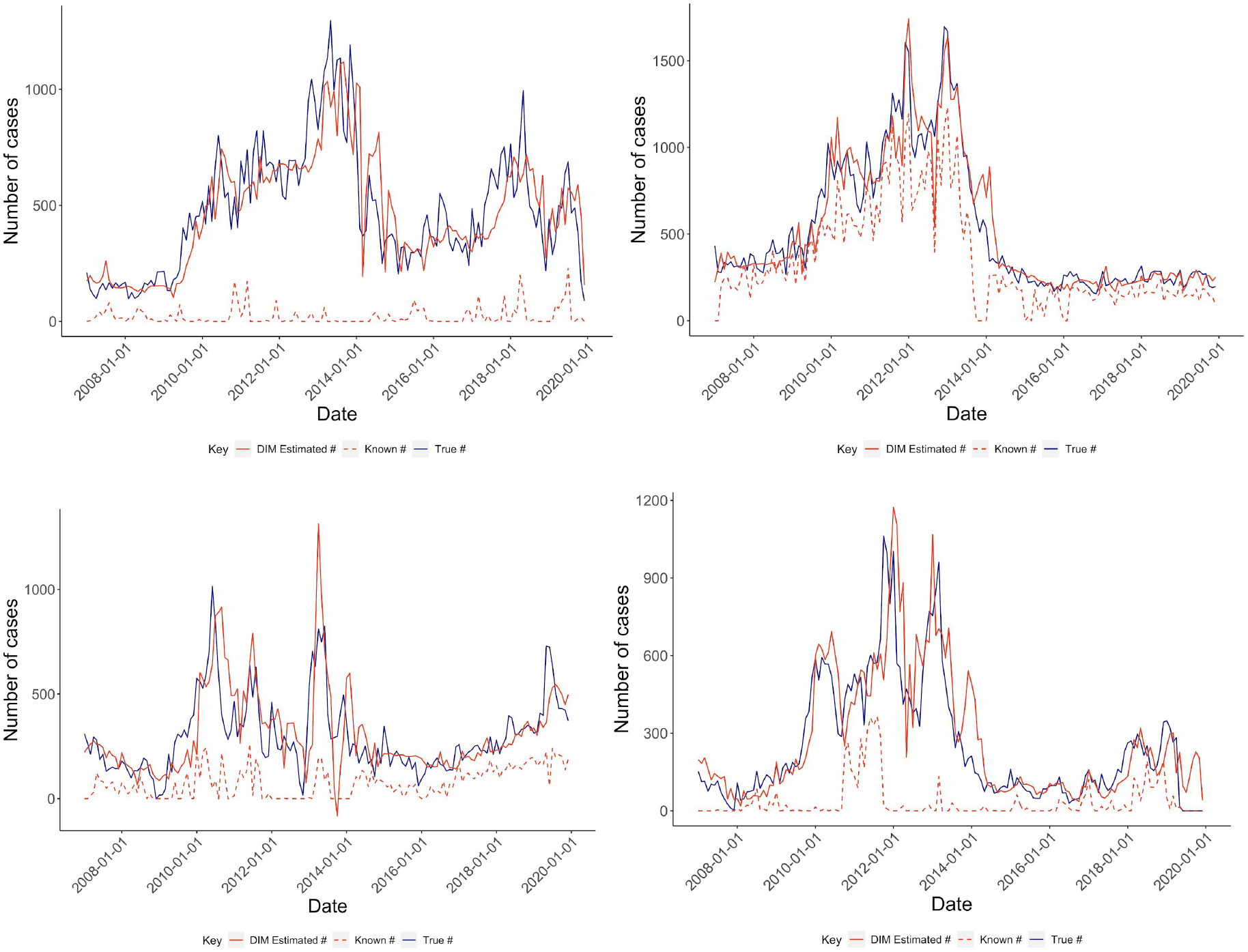
Results from Data Imputation Models for regions 1 (top left), 4 (top right), 7 (bottom left) and 8 (bottom right). Solid red lines indicate the number of cases estimated from the data imputation model, dashed red lines indicate the number of cases known by the end of the month and solid blue lines indicate the true number of eventually reported cases.

In contrast, the DIM models for regions 1, 7 and 8 significantly improved on revised case counts for each of these regions. Of these, the DIM model for region 8 reported the optimal performance (rRMSE= 0.127), exhibiting a slight lag in estimating revised cases (Figure 3). The DIM models for regions 1 and 7 reported weaker model performance (rRMSE=0.128 and 0.146, respectively), partially driven by the combined pronounced overestimation of revised cases in early 2013 and marked underestimation of revised cases in late 2013 and 2019, in the case of region 7, and the frequent underestimation of revised cases, in the case of region 1 (Figure 3). However, region 1 predictions broadly approximated true behavior from 2018 onwards, and model predictions for region 7 still generally reflected the most recent upward trend in cases from 2017, with the exception of late 2019, improving on month-end reporting substantially. Finally, the DIM model for region 9 failed to converge (identify a stable set of parameter values) due to data sparsity, largely driven by the exclusion of cases with implausible dates of documentation, as defined previously.

#### B. Network Models (NMs)

Both network models exhibited improvements for all regions (Figure 4). The NMs for region 1 resulted in predictions that more closely tracked revised case counts over time (Figure 4), showing reduced error rates for both models (rRMSE=0.116), compared to the data imputation models. NM2 predictions for region 1 reported only marginally greater accuracy than the corresponding predictions from NM1 (rRMSE=0.1157 vs 0.1164). The NMs for region 7 revealed perceptible gains in accuracy, more closely reflecting true dynamics in case counts and importantly, for both models, attenuating the appreciable drop and upswing in estimated cases in 2013 (Figure 4), with NM2 resulting in the greatest improvement (rRMSE=0.119 [NM2] and rRMSE=0.120 [NM1]). Finally, NM1 and NM2 improved over the DIM model for region 8 (rRMSE=0.1004 and 0.1001, respectively). Model predictions from 2013 onwards better mirrored the magnitude and timing of current trends, and partially corrected the significant overestimation of cases in late 2013 and late 2019 observed in the data imputation model predictions (Figure 4). As in regions 1 and 7, model predictions were most improved for NM2.

**Figure 4:**
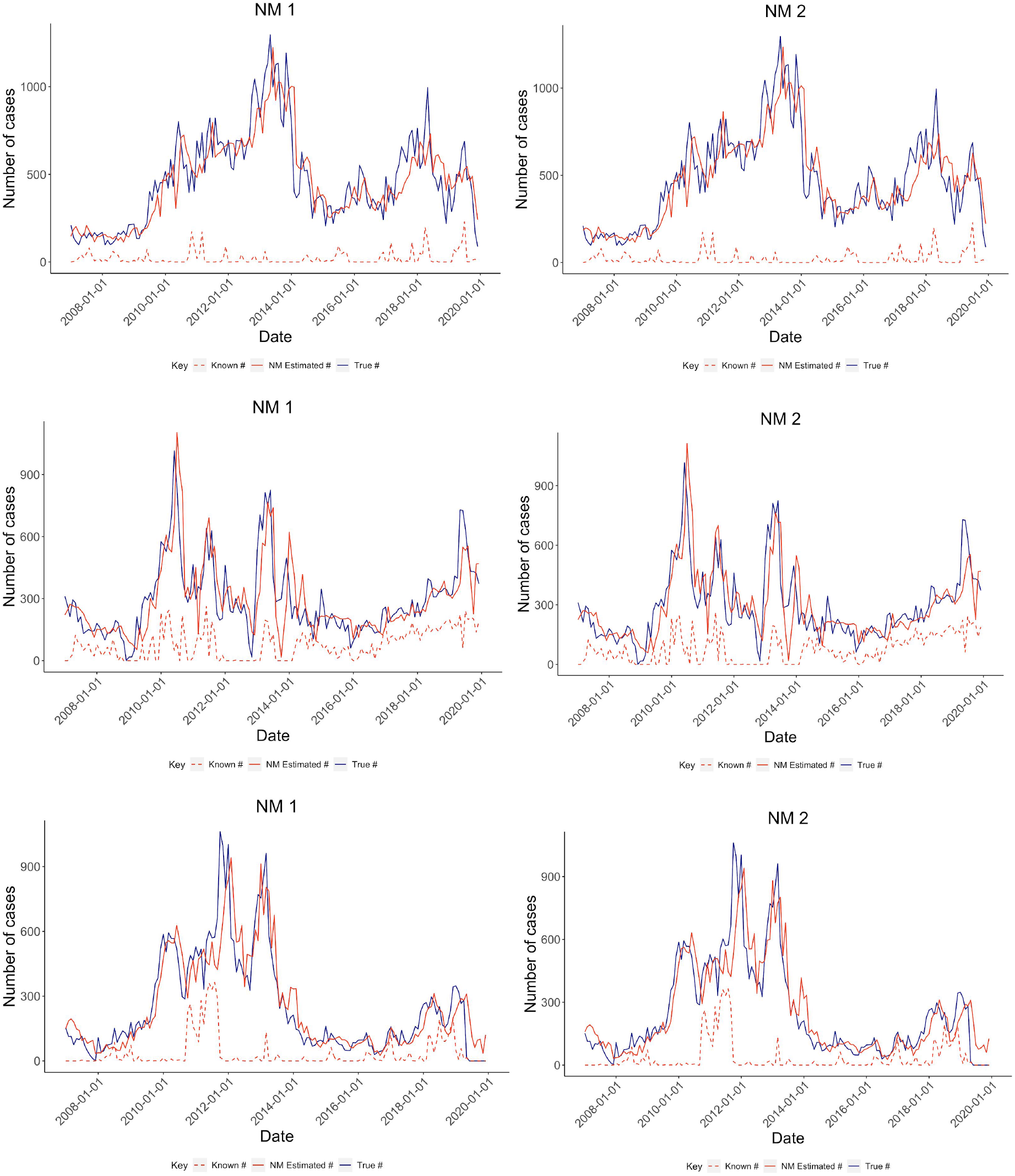
Results from Network models (NM1 and NM2) for regions 1 (top left and right), 7 (center left and right) and 8 (bottom left and right). Solid red lines indicate the number of cases estimated from the Network models, dashed red lines indicate the number of cases known by the end of the month and solid blue lines indicate the true number of eventually reported cases.

The data imputation and network models significantly improved over known case counts for regions 1, 4,7, and 8, most strikingly for regions 1 and 8, with error rates reduced by a factor of 3.8, 2.0, 2.3, and 2.9, respectively, for the regions’ best performing models (Supplementary Table S1). In all cases, when relying on predictions from the best performing models, the median fraction of cases captured within a month for all regions from 2017 to 2019 ranges from 0.96 to 0.99.

## Discussion

Our findings highlight the extent to which reporting lags pose a challenge to malaria surveillance in Guyana, with appreciable heterogeneities in the overall magnitude, spatial patterns, and time trends of delays, and potential social factors linked to observed delay patterns. While improvements to reporting systems, including digitization, infrastructure, and enhanced data management, are critical for malaria control in the medium- and long-term, many national programs will not have the resources to make significant changes in the near future. The nowcasting approach we describe here therefore represents a simple analytical method leveraging data that is often already being collected in order to significantly improve estimation of current malaria cases, at almost no increased investment. Our approach provides an easily accessible tool, which necessitates no additional data requirements and minimal training, and can be readily implemented in resource-constrained surveillance contexts.

We found convincing evidence for spatial clustering in delay patterns at the level of Neighborhood Democratic Councils, while synchronicities between reporting delays at the level of regions was more moderate. The minimal overlap of regional delay patterns may partially explain the modest improvements when incorporating time trends and predicted case counts in other regions to estimate region-specific case counts as done by the network models. Finally, we observed significant spatial associations between the presence of Amerindian settlements and mines and reporting delays, primarily occurring in regions 1, 7, 8 and 9, with the two overlapping in region 1. This finding suggests that issues in timeliness of reporting are most acutely distributed among socially remote and resource-limited communities.

In contrast, associations between precipitation and delays (from 2006-2019) were marginal. We note that this may be partially attributed to the fact that increased rains affecting driving conditions may undermine reporting timeliness at both a finer spatial scale than regions, or even neighborhood democratic councils, (i.e. with particular roads and towns threatened due to locally-specific infrastructural challenges) and at a finer temporal scale than months. Thus, these patterns are obscured in region- and monthly-level associations. Furthermore, the source of interpolated precipitation data may not fully represent real-world behavior, as “regions with sparse support [are affected by] patchy coverage”, as is the case in several areas within Guyana.^20^

Finally, the association between connectivity and delays in 2015 was minimal and non-significant. This finding can be explained by the significant variation in the level of connectivity -- both in terms of road access and other forms of travel -- observed within regions, which is again diluted in region-level associations.

The data imputation models reported relatively low error rates, suggesting that more simple models for each region that simply learn from their own autoregressive trends may be practical and straightforward to implement. While there was considerable variation in DIM model performance across regions, it is promising that the model achieved striking results for regions 1 and 8, where the lowest fraction of cases are reported within a month, and thus have the greatest need for improved real-time surveillance. Possible extensions to the network models to markedly improve their predictive power include accounting for other covariates that may better explain trends in reporting rates, such as access to waterways, another important source of travel. Additionally, the NMs may be best implemented using data on the previous distribution of known case counts by NDC (rather than by region), or at a more local spatial unit for which stronger relationships in reporting delays across locations are observed.

In addition to these limitations, there are a number of additional constraints of our modeling approach that should be addressed in future applications of our work and extensions to other settings. Our models do not allow flexible and non-linear relationships between the number of revised cases in a given region in month t and each the relevant predictors, such as potential interactions between the known case counts in month t-2 and month t-3, or between known case counts in month t-4 and precipitation (or other climate-related and other relevant covariates) in month t. Non-parametric methods like regression trees can instead be implemented to avoid these potentially erroneous model assumptions. Our approach can only be applied when there is sufficient data for model training and testing, as all models for region 9 failed to run due to sparse data, limiting its application to areas with sufficient malaria cases. However, despite these challenges, we believe that data imputation and network model approaches, which learn from time and spatial patterns in reporting, can greatly improve the current state of surveillance efforts, particularly for areas where data quality is poorest and remoteness lead to significant reporting delays.

## Data Availability

Raw data and code will be made available upon publication

## Supporting Information

**Supplementary Figure S1:**
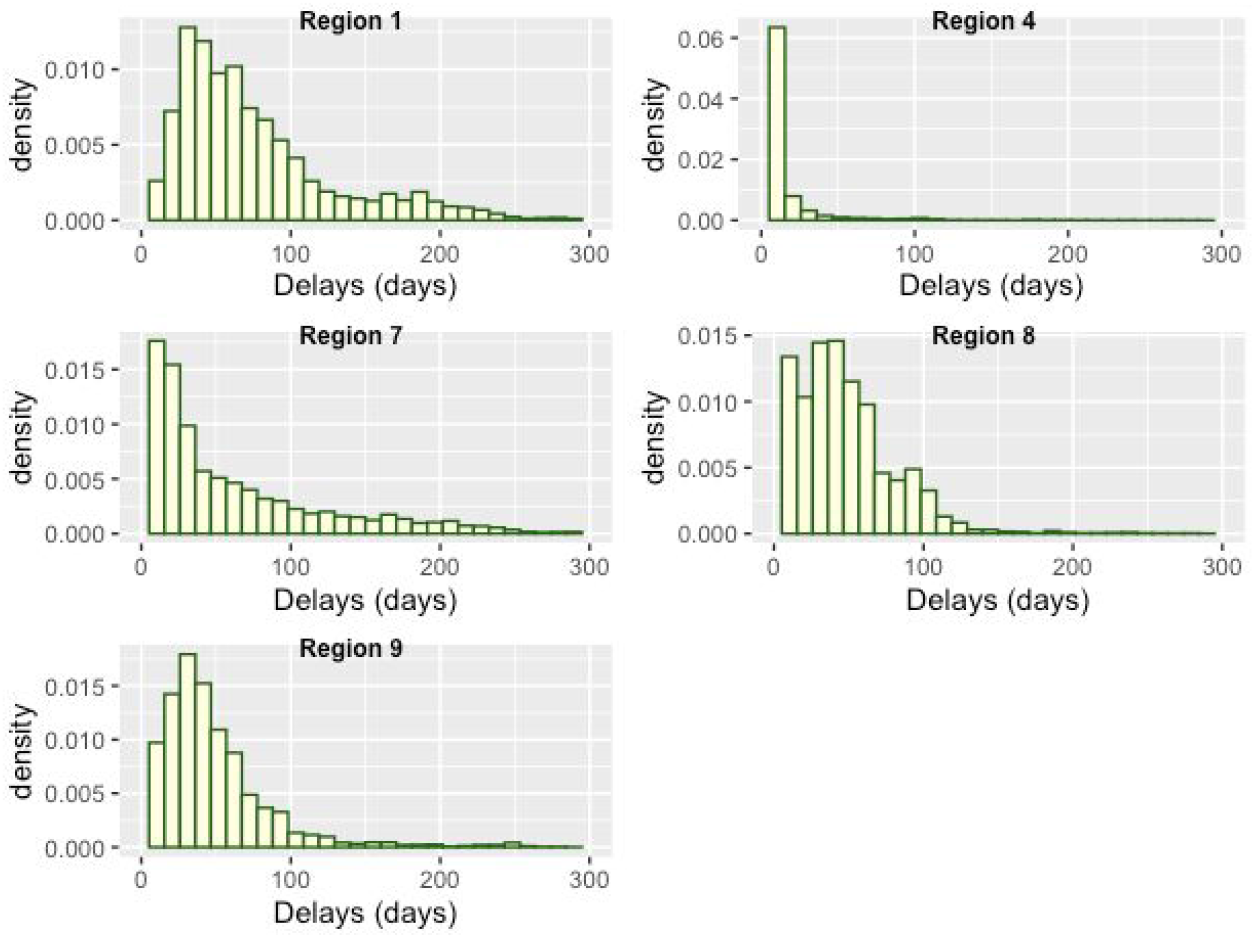
Distribution of delays from 2006 to 2019 for regions 1, 4, 7, 8 and 9.

**Supplementary Figure S2:**
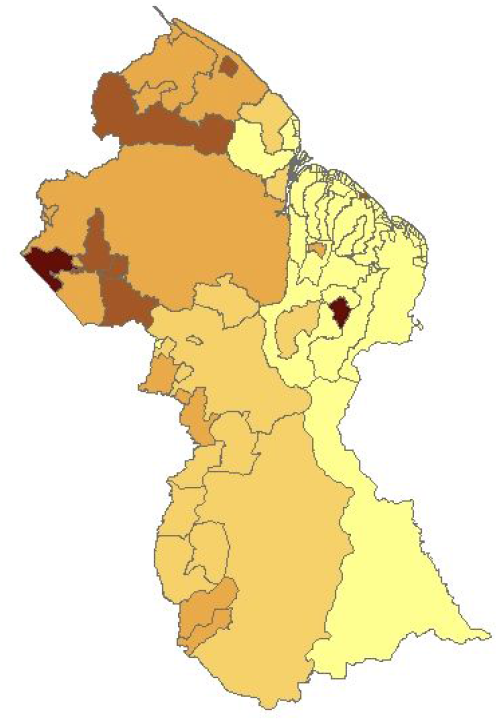
Map of median delays reported by region (darker colors indicate higher median delays)

**Supplementary Figure S3:**
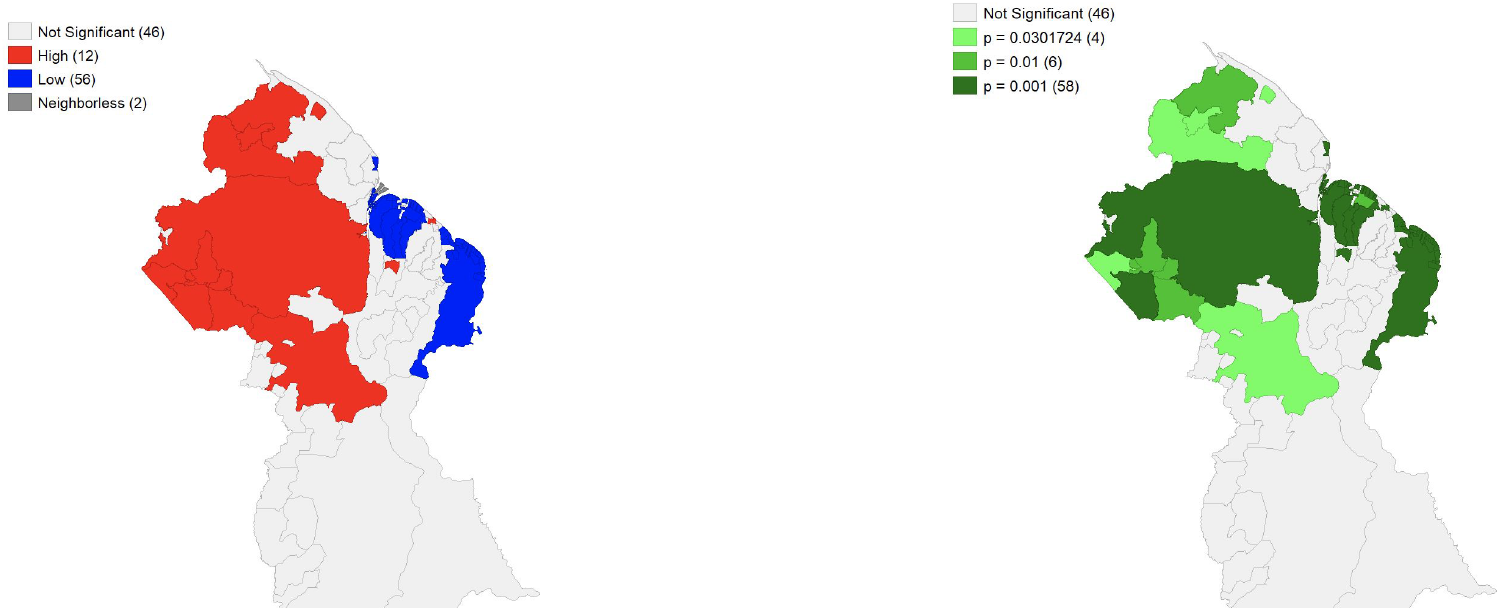
Local G* clustering maps of median delays (map of G* statistic (left) and FDR-adjusted significance map (right))

**Supplementary Figure S4:**
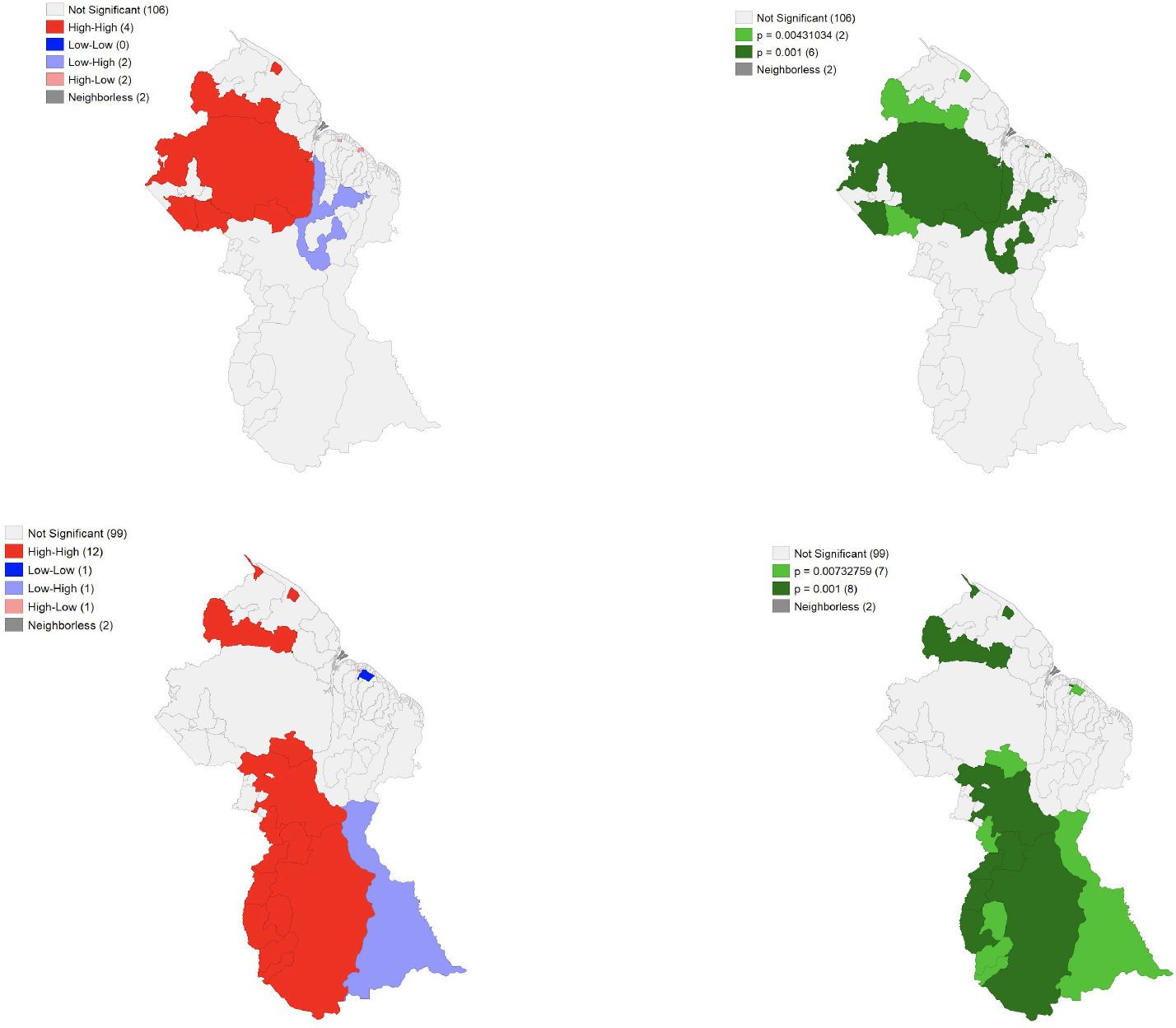
Local bivariate Moran’s I clustering maps of: aggregated median delays and density of mines (top left) and FDR-adjusted significance map (top right) and aggregated median delays and density of Amerindian settlements (bottom left) and FDR-adjusted significance map (bottom right).

**Supplementary Table S1:** Table of relative root mean squared errors (rRMSE) generated from data imputation models and network models for each region (note: for region 4, rRMSE only reported for the data imputation model, given that no network models were run for this region). The final row reports the best performing model* for each region, which corresponds to the model resulting in the lowest rRMSE.

|  | region 1 | region 4 | region 7 | region 8 |
| --- | --- | --- | --- | --- |
| DIM | 0.1280 | 0.0791 | 0.1464 | 0.1270 |
| NM 1 | 0.1164 | NA | 0.1199 | 0.1004 |
| NM 2 | 0.1157 | NA | 0.1187 | 0.1001 |
| Best performing model* | NM2 | DIM | NM2 | NM2 |

## Contributors

TFM, COB, MS, and HC designed the study. TFM performed the analyses, with input from CP, MS, SJW, and CC. TFM wrote the first draft of the manuscript, which was reviewed by all authors.

## Acknowledgments

We thank Ayesha Mahmud, Nishant Kishore, and Flavia Camponovo for their valuable suggestions and support and Marcia Castro and Jack Cordes for helpful discussions related to the spatial analyses. We are indebted to the entire VCS team in the central office and the Bartica District Hospital for all their work in facilitating this data collection effort and for detailing and contextualizing the process of reporting.

## Note on the Data Source

This dataset was obtained for public health purposes and reanalyzed for research under agreement with the Guyana Ministry of Public Health.

